# Returning to school during Ebola: an exploratory health-zone framework for scenario-based school-system introduction pressure during the September 2026 rentrée in eastern Democratic Republic of the Congo

**DOI:** 10.64898/2026.08.20.26360976

**Authors:** Johan G. L. Verheyden, Celestin Nzanzu Mudogo

## Abstract

School reopening during an Ebola outbreak is often framed as a binary question of whether schools are safe. For Ebola, however, the immediate operational question is where an infected school-age child may reach the school system before recognition and isolation, and whether local systems can detect and respond rapidly. We developed an exploratory, scenario-based health-zone framework for the September 2026 rentrée during the ongoing Bundibugyo virus disease outbreak in eastern Democratic Republic of the Congo. The primary estimand was scenario-based expected introduction pressure, expressed on an expected-count scale, for infected school-age children reaching school in each health zone during a one-week window. The framework combined recent reported transmission, estimated school-age exposure and attendance, a surveillance/interception probability, and directed mobility-based importation. Case-fatality patterns were analysed separately and did not determine introduction pressure. A 10,000-draw probabilistic sensitivity analysis examined uncertainty in the school-age case share, attendance, pre-isolation school-entry probability and mobility scaling. Geographic components were retrospectively evaluated at eight non-overlapping weekly origins from 1 June to 20 July 2026, using subsequent reported seven-day health-zone activity and first reported cases in previously unaffected zones as outcomes. Seven-day local epidemic pressure discriminated health zones with subsequent reported activity with pooled ROC-AUC 0.848; the 14-day local measure increased this to 0.885. Adding directed mobility increased ROC-AUC to 0.952. In the base scenario, the six-province combined scenario-based expected introduction pressure was 10.97; the probabilistic sensitivity median was 11.17, with a 2.5–97.5% sensitivity range of 5.56–20.97. Bunia, Rwampara and Nizi had the highest base introduction pressures, followed by Katwa and Nia Nia. Among 24 previously unaffected health zones that subsequently reported a first confirmed case, 13 (54.2%) were in the top 10 and 18 (75.0%) in the top 20 mobility-ranked zones; random selection would have been expected to capture approximately 2.05 and 4.10 events, respectively. In a separate six-origin exploratory nested-specification sensitivity, surveillance/access modifiers did not improve geographic discrimination over local epidemic pressure alone, whereas mobility did. The dominant structural uncertainty remained the probability that an infected child reaches school before being identified. The framework supports targeted geographic prioritisation and minimum school-health readiness, but its probabilities are model-implied scenario probabilities rather than calibrated forecasts or evidence for a single national open/close decision.

## Introduction

The September 2026 return to school in the Democratic Republic of the Congo (DRC) will occur during an active Bundibugyo virus disease (BVD) outbreak in the east of the country, which remained characterised by sustained transmission and geographic expansion during July 2026 [1]. The official 2026–2027 school calendar places the formal rentrée on 1 September 2026 [2], but registration, teacher mobilisation, school preparation and household movement begin before the first day of classes. The relevant exposure period is therefore not a single morning: it is a short period in which pupils, teachers and caregivers resume regular journeys between homes, schools, transport routes, markets, churches and other community settings.

Ebola creates a different school-health problem from airborne respiratory epidemics. Human-to-human transmission occurs principally through direct or close contact with symptomatic infected persons, body fluids or contaminated materials [3,4]. In a school, ordinary co-presence in a classroom is therefore not the main mechanism of concern. More plausible pathways include an ill pupil or teacher attending before recognition, classmates or staff helping a person who vomits or becomes acutely unwell, contact with blood or other body fluids, unsafe cleaning, delayed referral, and concealment of symptoms or exposure because of fear or stigma. Joint WHO, UNICEF and CDC guidance developed during the West African Ebola epidemic accordingly emphasised illness recognition, keeping sick pupils and staff away from school, hand hygiene, safe cleaning, reduced direct physical contact, communication and rapid linkage to health services [5].

The literature also cautions against treating schools as automatic Ebola amplifiers. Schools are organised contact settings, but they can also be organised detection and communication settings. During the 2018–2020 eastern DRC Ebola response, UNICEF and education authorities mapped high-risk schools, installed handwashing facilities and thermometers, supplied hygiene materials, trained teachers and administrators, adapted messages using teacher feedback, and strengthened outreach to parents [6,7]. Teachers and pupils were used not only as recipients of prevention messages but as trusted channels through which information could move back into households. This precedent supports a concept of minimum school-health readiness rather than an assumption that every school can or must become a fully equipped health facility.

That distinction matters because the physical and organisational conditions of Congolese schools are highly heterogeneous. Following the introduction of free public primary education, nearly four million additional pupils were enrolled in public primary schools in 2022–2023 compared with 2017–2018, and the World Bank has identified overcrowded classrooms, shortages of teaching resources, and access to water and sanitation as continuing constraints [8]. In eastern DRC, conflict and displacement add another layer. Schools may lose classrooms to damage, be used temporarily as shelters, absorb displaced pupils, or operate through temporary learning spaces. In one documented school in Minova, 600 pupils were enrolled in six classrooms before an additional 137 displaced children were expected for the new school year [9]. In other locations, temporary classrooms have been constructed rapidly from locally available materials because existing schools cannot accommodate displaced and host-community children [10].

There is therefore no single ‘DRC classroom’ to which one uniform school-risk assumption can be applied. In dense urban and peri-urban settings, the operational challenge may be the number of pupils moving through crowded classrooms, entrances, courtyards and transport routes. In rural or remote settings, smaller formal enrolment does not necessarily imply easier control: water and sanitation may be less reliable, buildings may be more fragile, temporary learning spaces more common, and referral to a health facility or testing site may require longer travel. As an older national structural benchmark, MICS 2018 indicators cited by UNICEF and Education Cannot Wait reported that only 38% of primary schools had access to water and that primary-school completion was 48% in rural areas compared with 79% in urban areas [11].

These figures should not be interpreted as current health-zone measurements, but they illustrate longstanding inequalities in the conditions under which national prevention guidance must be implemented. Conflict-affected areas can combine both patterns—crowding caused by displacement with weak infrastructure and disrupted access. These are stylised operational contrasts rather than a binary urban–rural classification, but they are important because the consequences of one missed introduction depend on what a school can actually do once illness is recognised [8–14].

Water, sanitation and hygiene are particularly relevant because Ebola school safety depends on the ability to wash hands, clean body-fluid contamination and manage an ill person without exposing other pupils or staff. Previous UNICEF work in DRC found that many schools had access to at least one water, sanitation or hygiene component but rarely all components required for effective infection prevention and control [14]. This does not mean that schools without complete WASH infrastructure inevitably amplify Ebola; it means that a recommendation such as ‘wash hands immediately’ or ‘clean contaminated surfaces safely’ can be operationally very different across schools.

The 2026 education response already recognises schools as part of the national Ebola prevention system. In May 2026, the Ministry of Education instructed schools and examination centres to reinforce handwashing, immediately report suspected cases and reorganise spaces to reduce close contact; the President subsequently directed intensified sensitisation in schools and universities [15,16]. These instructions provide a national prevention benchmark, but a national instruction is not evidence that every school has the water, space, staff confidence or referral linkage required to implement it. The analytical problem is therefore partly epidemiological and partly operational.

School closure is also not a neutral default. Evidence from Ebola and other epidemics links prolonged closure with disrupted enrolment, child labour, early marriage, adolescent pregnancy, exploitation and loss of protective routines [17,18]. Eastern DRC experience is especially relevant because insecurity and displacement already keep large numbers of children out of school: by May 2025, UNICEF reported more than 1.3 million out-of-school children in Ituri alone, with hundreds of schools damaged or destroyed [12]. At the same time, recent education-resilience programmes in Ituri explicitly frame safe education as part of child protection and recovery in crisis settings [13]. A realistic analysis must therefore compare the risks of school attendance with the risks and practical consequences of displacement from school, rather than assuming that closure simply removes exposure.

Against this background, the research gap is not whether Ebola can occur in a school, nor whether all schools should open or close. The more useful question is where existing community transmission, population movement and imperfect interception make the arrival of an infected school-age child sufficiently plausible to require additional attention. We therefore developed an exploratory health-zone framework that estimates scenario-based school-system introduction pressure, separates local and imported pressure, quantifies uncertainty, retrospectively evaluates the geographic components of the model, and interprets the numerical results against the contemporary operational environment.

### Study objective

The primary objective was to estimate health-zone-level BVD introduction pressure into the school system during the September 2026 rentrée and identify where intensified monitoring or targeted mitigation may be most justified. The analysis was not intended to support a single national open/close decision, to predict transmission within individual schools, or to estimate deaths attributable to school attendance. A secondary objective was to test whether the geographic components of the framework—recent epidemic pressure and mobility—could retrospectively distinguish health zones that subsequently reported activity or first cases.

## Materials and methods

### Study design and analytical unit

We conducted a health-zone-level risk-estimation study combining recent outbreak surveillance, modelled population and school exposure, response-capacity proxies, directed population mobility and structured documentary contextualisation. The health zone was the primary analytical unit because it is the operational level at which recent incidence, contact tracing, access to testing and response activity vary substantially. The quantitative study area comprised 151 health zones across six eastern outbreak-relevant provinces included in the analytical dataset.

### Data sources

Epidemiological data were drawn from DRC national SitReps and the INRB-UMIE BDBV2026-Data repository, which manually transcribes SitRep indicators and integrates contextual public-health response data [19,20]. Health-zone population denominators were taken from the WorldPop layer contained in the repository. School-age population and attendance were estimated from UNICEF demographic and education indicators when health-zone-specific denominators were unavailable [21]. Directed mobility was represented using Flowminder origin–destination data and IOM displacement flows contained in the repository [22,23]. All variables retained provenance and were labelled as observed, reconstructed, interpolated or extrapolated. The model was frozen at the 11 August 2026 data cut-off. Because the formal rentrée was scheduled for 1 September 2026, and because administrative mobilisation and actual return-to-class timing may vary locally, the analysis should be interpreted as a pre-rentrée operational snapshot rather than a direct observation of September school attendance. A final refresh closer to the rentrée can update inputs without changing the model specification. To interpret the quantitative findings against the response environment immediately preceding the snapshot, we reviewed the ten most recent available national SitRep sources by report date (31 July–11 August 2026) [20]. Operational notes were treated as contemporaneous snapshots rather than a continuous qualitative series; non-mention of a problem was not interpreted as evidence of absence, and no contextual theme was converted into a numerical model weight.

The detailed coding matrix is provided in the supplementary material. Table 1 summarises the analytical role, evidence base and validation status of each major model component.

**Table 1.** Model components, analytical role and validation status.

| Component | Operational quantity / input | Role in model | Validation status | Main caution |
| --- | --- | --- | --- | --- |
| Recent epidemic pressure | Recent 7-day HZ case increment; 14-day weekly-equivalent sensitivity | Defines local/source infection pressure | 7d pooled ROC-AUC 0.848; 14d 0.885 | Reported pressure, not latent incidence. |
| School-age case share | 5–17 share from SitRep 082; base 13.17%, sensitivity 10–16% | Scales cases to school-age infections | Scenario sensitivity | National extrapolation, not HZ-specific. |
| Attendance | Base 56.6%; sensitivity 45–70% | Scales school-age infections to expected attendance | Scenario sensitivity | Not observed rentrée attendance. |
| School-entry/interception $q$ | Base 0.25; sensitivity 0.10–0.50 with HZ proxy modifiers | Converts infected pupils to local $\lambda$; applies at destination for imported $\lambda$ | Not directly externally calibrated | Dominant structural uncertainty; not an observed failure rate. |
| Directed mobility | Flowminder OD + IOM DTM | Distributes source pressure to destination HZs | Mobility ROC-AUC 0.952; first-HZ ROC-AUC 0.876 | Relative/scenario flows; absent/redacted links can create false reassurance. |
| CFR / severity | Shrunk HZ fatality residuals | Not included in primary $\lambda$ | Secondary sensitivity only | Confounded by delay, ascertainment, age mix and reporting. |
| Recent SitRep context | Structured review of latest 10 available reports | Interpretation and triangulation only | Qualitative contextualisation | Non-reporting is not evidence of absence; no numerical weight. |

### School-age population and attendance

In the absence of a complete harmonised health-zone school census, the population aged 5–17 years was estimated using a national age-structure share of 31.96%, derived from UNICEF/UN population age bands [21]. Expected school attendance was represented with a base value of 56.6%, obtained by age-weighting UNICEF education indicators used in the analytical workbook (43% at age 5, 78% for primary-age children, 32% for lower-secondary age and 34% for upper-secondary age); low and high sensitivity values were 45% and 70% [21]. The proportion of confirmed cases occurring among persons aged 5–17 years was based on the latest usable standardised SitRep demographic snapshot (455 of 3,454 cases; 13.17%) and was varied from 10% to 16% in sensitivity analyses [20]. These quantities are exposure assumptions rather than observed attendance on 1 September and were therefore propagated as uncertain inputs.

### Primary estimand and local introduction model

The primary estimand, λ_h,T_, was the scenario-based expected introduction pressure, expressed on an expected-count scale, for infected school-age children reaching the school system in health zone *h* during time window *T*. Recent seven-day reported case incidence was used as the principal epidemic-pressure signal. A simple one-week continuation assumption projected the next school-week case burden from the most recent reconstructed seven-day case count; a weekly-equivalent 14-day pressure measure was examined as a robustness sensitivity.

For each health zone, projected cases were multiplied by the assumed 5–17-year case share and expected attendance to obtain the expected number of infected pupils who would otherwise attend. This quantity was then multiplied by q_h_, the probability that an infected child reaches school before recognition, isolation or exclusion from attendance. The mapping from λ_h,T_ to 1 − exp(−λ_h,T_) is a conditional Poisson-process transformation used to express expected pressure as a model-implied scenario probability. It is not an empirically calibrated probability forecast, and the Poisson assumption is not tested for clustered household, mobility or school-linked events (Fig 1).

**Fig 1.**
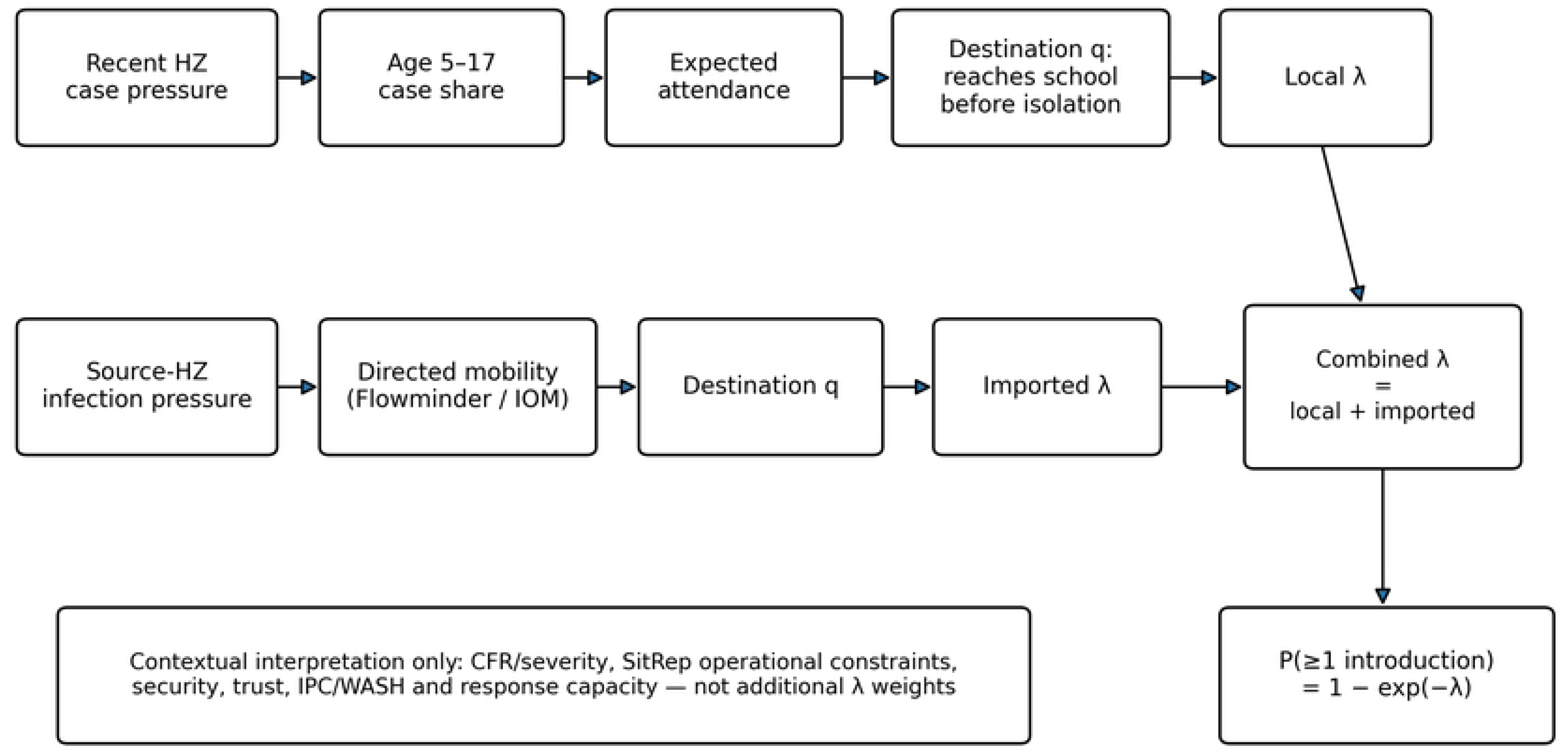
Conceptual framework. Recent health-zone epidemic pressure is scaled to school-age infections and expected attendance, then combined with the probability q that an infected child reaches school before recognition or isolation. Imported pressure is distributed through directed mobility and adjusted by destination q. CFR, SitRep operational context and other response constraints are interpreted separately rather than converted into additional λ weights.

### Surveillance/interception adjustment

The base pre-isolation school-entry probability q was set to 0.25, with low and high scenarios of 0.10 and 0.50. Health-zone q values were adjusted using analyst-specified structural modifiers for contact follow-up, testing access, health-site density, documented school-response activity and data quality: q_h_ = clip(q_0_ × m_contact,h_ × m_PCR,h_ × m_site,h_ × m_school,h_ × m_data,h_, 0.05, 0.75). The modifier directions were specified a priori: weaker contact follow-up, longer modelled PCR access and poorer data quality increase q; greater health-site density and documented school-response activity reduce q. These weights are not fitted coefficients. Contact tracing was treated as part of the interception process rather than as a transmission-rate covariate, reflecting the role of monitoring and early isolation of contacts in Ebola control [24]. Because contact-follow-up indicators are affected by reporting completeness, the resulting q values should be interpreted as transparent scenario adjustments, not directly observed probabilities.

### Mobility and imported introduction pressure

Total scenario-based expected introduction pressure was separated into local and imported components. Origin health-zone infection pressure was defined before applying the surveillance-interception probability. For destination *h*, imported pressure was calculated as λ_imp,h_ = q_h_ [f_F_ Σ_o≠h_ A_o_ W^F^_oh_ + f_I_ Σ_o≠h_ A_o_ W^I^_oh_], where A_o_ is origin infected school-age attendance pressure before interception, wFoh and wIoh are Flowminder and IOM origin–destination shares, and f_F_ and f_I_ are scenario movement fractions. Self-flows were removed. For each origin, available non-self OD entries were normalized across the destinations represented in the study matrix; origins with no usable row contributed no imported pressure. The destination health zone’s q adjustment was then applied. This separates infection pressure at the origin from detection/interception capacity at the destination.

### CFR as a secondary severity/capacity sensitivity

Crude case-fatality ratios were not used as transmission drivers. Health-zone fatality patterns are affected by outcome delay, under-ascertainment, age mix, referral patterns and reporting completeness [26,27]. We therefore analysed CFR separately using empirical-Bayes shrinkage toward a leave-one-out provincial comparator, with a prior effective sample size of 20. The resulting residuals were treated only as descriptive severity/response-capacity signals and did not modify λ_h,T_ or the primary health-zone ranking.

### Probabilistic sensitivity analysis

We conducted 10,000 Monte Carlo draws with a fixed random seed. Structural assumptions for the 5–17 case share, school attendance, baseline q, Flowminder movement fraction and IOM displacement fraction were drawn from split-triangular distributions with the base value as the median. Week-to-week case-count variability was represented by a Poisson draw with mean equal to the most recent seven-day case count. This Poisson step is a simple process-variation device and does not establish that introductions are independently Poisson in reality. The resulting 2.5th–97.5th percentile ranges are sensitivity intervals: they combine uncertainty under specified assumptions and simple process variation, and should not be interpreted as calibrated confidence or predictive intervals.

### Retrospective component validation

We retrospectively evaluated the geographic components of the framework using eight non-overlapping weekly cut-offs: 1 June, 8 June, 15 June, 22 June, 29 June, 6 July, 13 July and 20 July 2026. For each origin, only information available on or before the cut-off was used to generate model scores, and subsequent reported health-zone activity was assessed over the following seven days. The primary validation therefore ends on 27 July, before the later health-zone series becomes less complete.

Health-zone case histories were reconstructed from the reconciled direct-PDF series rather than from a single repository cumulative-case file. During validation we found that the repository’s main health-zone cumulative-case CSV becomes stale for some established zones after changes in SitRep reporting format, whereas the reconciled dataset contains later direct-PDF health-zone extractions. Using the reconciled series avoids counting transcription gaps as epidemiological non-events.

Two outcomes were considered. First, future health-zone activity was defined as any reported increase in cumulative confirmed cases during the subsequent seven days. Second, geographic spread was defined as the first reported confirmed case in a health zone that had not yet reported a case at the cut-off. Recent epidemic pressure was reconstructed over approximately seven and fourteen days using the closest available observations, with the fourteen-day increment expressed as a weekly-equivalent rate. Negative revisions were not treated as negative epidemic pressure.

As a secondary exploratory sensitivity, we compared three nested specifications: local epidemic pressure alone; local pressure with surveillance and access modifiers; and the same specification with directed mobility. This comparison used six overlapping validation origins (13, 15, 17, 18, 19 and 20 July 2026) for which all three specifications had been reconstructed using information available at or before each cut-off. The six-origin sample is therefore different from the primary eight-origin validation, which used non-overlapping weekly cut-offs (1, 8, 15, 22 and 29 June; 6, 13 and 20 July). Because the six origins overlap and are concentrated in a later epidemic phase, their pooled discrimination metrics are not directly comparable with the primary eight-origin ROC-AUC values; they are used only to compare the three nested specifications within the same six-origin sample. Static PCR-access and health-site modifiers were retained; school-response evidence was used only when dated on or before the cut-off; contemporaneous or latest available contact-follow-up information was used; and final case-series quality labels were excluded because they would incorporate future information.

Discrimination was summarized using ROC-AUC and average precision; retrieval of subsequently active or newly affected health zones was assessed using top-10 and top-20 capture; and Spearman rank correlation was used to compare model scores with the magnitude of subsequent reported case increments. Because health zones recur across rolling origins, and because the six secondary nested-specification origins have overlapping seven-day outcome windows, pooled metrics are interpreted descriptively rather than as independent inferential observations. We therefore added origin-specific distributions and an origin-block bootstrap over supplied origin summaries as sensitivity diagnostics, while avoiding inferential claims from the small number of origins. For first-HZ spread, we compared top-10/top-20 capture and AP with prevalence-aware random-selection baselines. This validation assesses future reported activity and geographic model components, not the exact probability q that an infected pupil reaches school before recognition or isolation.

### Software and reproducibility

Analyses and reproducibility checks were implemented in Python 3.13.5. Numerical operations used NumPy 2.3.5; retrospective discrimination metrics used scikit-learn 1.8.0; and figures were generated with Matplotlib 3.10.8. The PSA and deterministic q-scenario calculations are exactly reproducible from S2 File, including the random seed, raw 10,000 draws and mobility matrices. The validation hardening outputs—origin distributions, origin-block bootstrap summaries and first-HZ random baselines—are reproducible from the supplied origin/event summaries. The package does not fully reconstruct the original rolling-origin score matrices from dated health-zone case-count histories; this boundary is stated in S2 rather than implied away.

### Ethics statement

This methodological study used publicly available, aggregated and de-identified outbreak surveillance information and public historical archives and did not involve recruitment, intervention, or access to identifiable individual-level records. Formal ethics committee review and participant consent were therefore not required.

### Interpretation and scope

A high λ indicates greater scenario-based school-system introduction pressure, not that a particular school will receive a case or that school attendance will cause transmission. Likewise, 1 − exp(−λ) is a model-implied scenario probability for the health-zone school system and must not be interpreted as a calibrated individual-school risk. The framework estimates encounter pressure at health-zone level; it does not estimate secondary transmission within schools.

## Results

### Overall introduction pressure

At the 11 August cut-off, the preferred base specification produced a combined scenario-based expected count of 10.97 school-system introductions across the six-province analytical scope during the modelled week. This number is an expectation under the model assumptions, not a forecast that exactly eleven infected pupils will attend school during September. Repeated weeks under the same underlying conditions would not produce the same realised count each time; λ summarises the average pressure implied by the inputs. Because the cut-off precedes the formal 1 September rentrée by about three weeks, the result should be read as an operational pre-rentrée snapshot rather than a direct September forecast.

The expected burden was geographically concentrated. Ituri accounted for approximately 8.47 of the 10.97 combined expected introductions, or about three quarters of the total modelled pressure. North Kivu contributed about 1.39 and Haut-Uélé about 1.03, while Tshopo, South Kivu and Bas-Uélé contributed much smaller absolute expectations. This concentration reflects the location of recent reported transmission; it should not be interpreted as evidence that lower-burden provinces are risk-free.

### Health-zone ranking

Bunia had the highest base introduction expectation (λ=2.08; P≥1=87.4%), followed by Rwampara (λ=1.84; 84.1%) and Nizi (λ=1.32; 73.4%). Katwa in North Kivu and Nia Nia in Ituri each had a modelled probability of at least one introduction of approximately 50%. For a non-statistical interpretation, Bunia’s λ of about 2 does not mean that two specific children are predicted to attend school infected; it means that the combination of recent case pressure, the estimated school-age share, attendance, interception assumptions and mobility produces an expected count close to two for the health-zone school system as a whole. Likewise, P≥1 is a health-zone school-system probability, not the probability for an individual school (Table 2; Fig 2).

**Fig 2.**
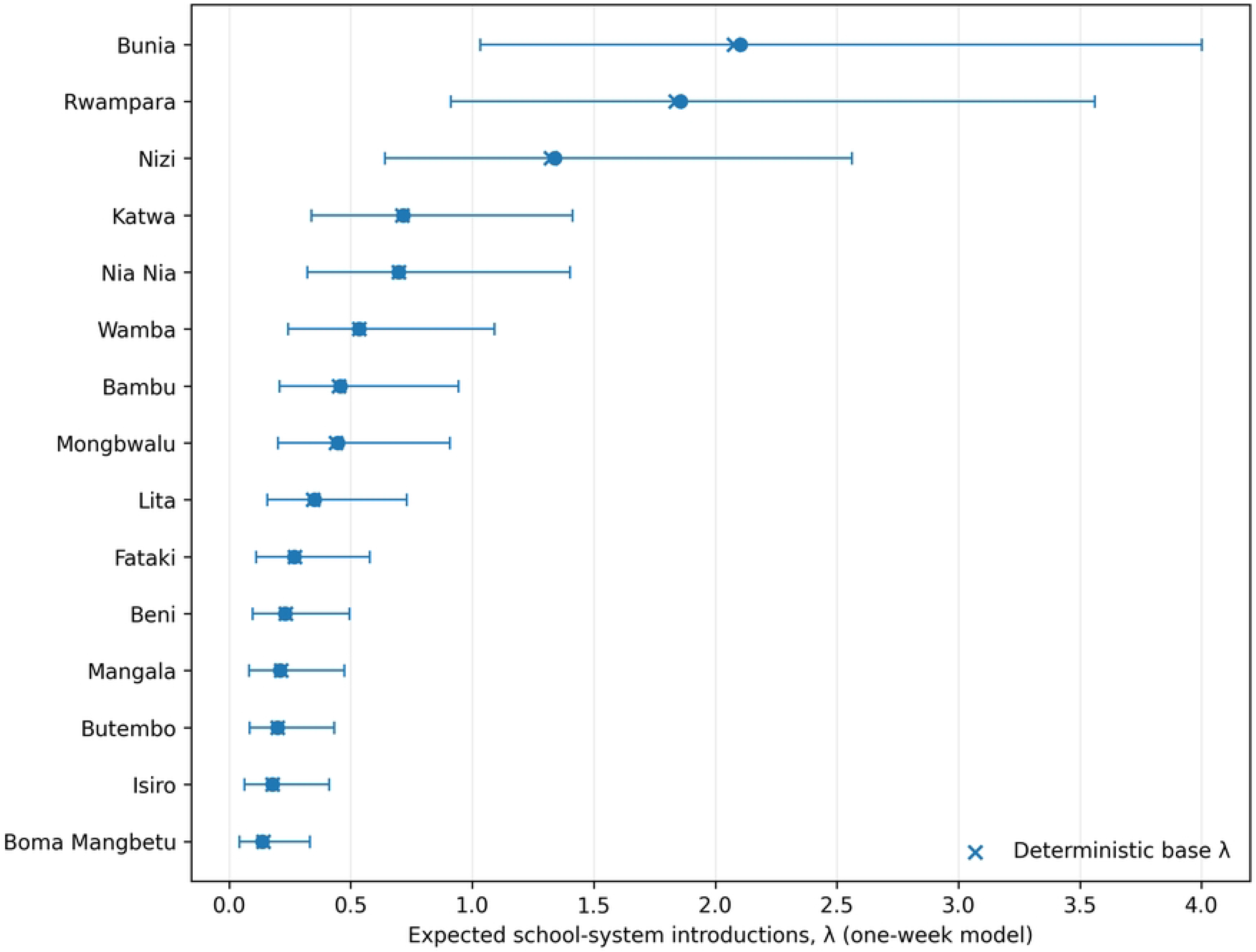
Deterministic base λ and probabilistic sensitivity intervals for the 15 highest-ranked health zones. Circles show PSA medians with 2.5–97.5 percentile sensitivity intervals; crosses show deterministic base estimates. Intervals combine structured uncertainty in model assumptions with simple week-to-week case-count variation and are not calibrated confidence intervals.

**Table 2.** Highest modelled school-system introduction pressure at the 11 August 2026 cut-off.

| Rank | Health zone | Province | Combined $\lambda$ | $P(\geq 1)$ | PSA $\lambda$ median (2.5–97.5%) | $P(\text{top } 10)$ | 14d rank |
| --- | --- | --- | --- | --- | --- | --- | --- |
| 1 | Bunia | Ituri | 2.075 | 87.4% | 2.103 (1.033–4.001) | 100.0% | 1 |
| 2 | Rwampara | Ituri | 1.837 | 84.1% | 1.857 (0.912–3.562) | 100.0% | 2 |
| 3 | Nizi | Ituri | 1.324 | 73.4% | 1.340 (0.641–2.562) | 100.0% | 3 |
| 4 | Katwa | Nord-Kivu | 0.713 | 51.0% | 0.717 (0.338–1.414) | 100.0% | 4 |
| 5 | Nia Nia | Ituri | 0.697 | 50.2% | 0.698 (0.322–1.403) | 100.0% | 5 |
| 6 | Wamba | Haut-Uele | 0.536 | 41.5% | 0.534 (0.244–1.092) | 99.8% | 10 |
| 7 | Bambu | Ituri | 0.452 | 36.4% | 0.457 (0.208–0.944) | 99.5% | 8 |
| 8 | Mongbwalu | Ituri | 0.439 | 35.5% | 0.447 (0.202–0.908) | 99.0% | 6 |

| Rank | Health zone | Province | Combined $\lambda$ | P($\geq 1$) | PSA $\lambda$ median (2.5–97.5%) | P(top 10) | 14d rank |
| --- | --- | --- | --- | --- | --- | --- | --- |
| 9 | Lita | Ituri | 0.346 | 29.2% | 0.352 (0.157–0.731) | 91.3% | 7 |
| 10 | Fataki | Ituri | 0.270 | 23.7% | 0.269 (0.111–0.578) | 53.2% | 9 |
| 11 | Beni | Nord-Kivu | 0.232 | 20.7% | 0.230 (0.097–0.495) | 23.0% | 11 |
| 12 | Mangala | Ituri | 0.213 | 19.2% | 0.209 (0.082–0.474) | 15.9% | 14 |
| 13 | Butembo | Nord-Kivu | 0.199 | 18.0% | 0.200 (0.085–0.433) | 8.8% | 12 |
| 14 | Isiro | Haut-Uele | 0.179 | 16.3% | 0.178 (0.064–0.412) | 6.2% | 13 |
| 15 | Boma Mangbetu | Haut-Uele | 0.140 | 13.0% | 0.136 (0.043–0.332) | 1.1% | 17 |

Note: $\lambda$ is an expected health-zone school-system count. PSA limits are sensitivity percentiles under specified assumptions, not calibrated confidence intervals.

The ranking is more informative than the small differences between adjacent point estimates. Bunia, Rwampara and Nizi form a clearly higher-risk cluster, while Katwa, Nia Nia, Wamba, Bambu, Mongbwalu and Lita remain consistently elevated across uncertainty analyses. This matters operationally because prioritisation decisions can be reasonably stable even when the exact expected count is not.

### Effect of mobility

Mobility increased the geographic reach of the model more than it changed the highest-risk ordering. Sixty-eight health zones with no recent local cases acquired a non-zero imported component in the base mobility scenario, while all ten highest local-risk health zones remained in the top ten after mobility was added. In other words, movement does not erase the importance of local transmission; it prevents the model from treating a connected health zone as epidemiologically isolated simply because no recent local case was reported. The composition of local and imported pressure among the highest-ranked health zones is shown in Fig 3.

**Fig 3.**
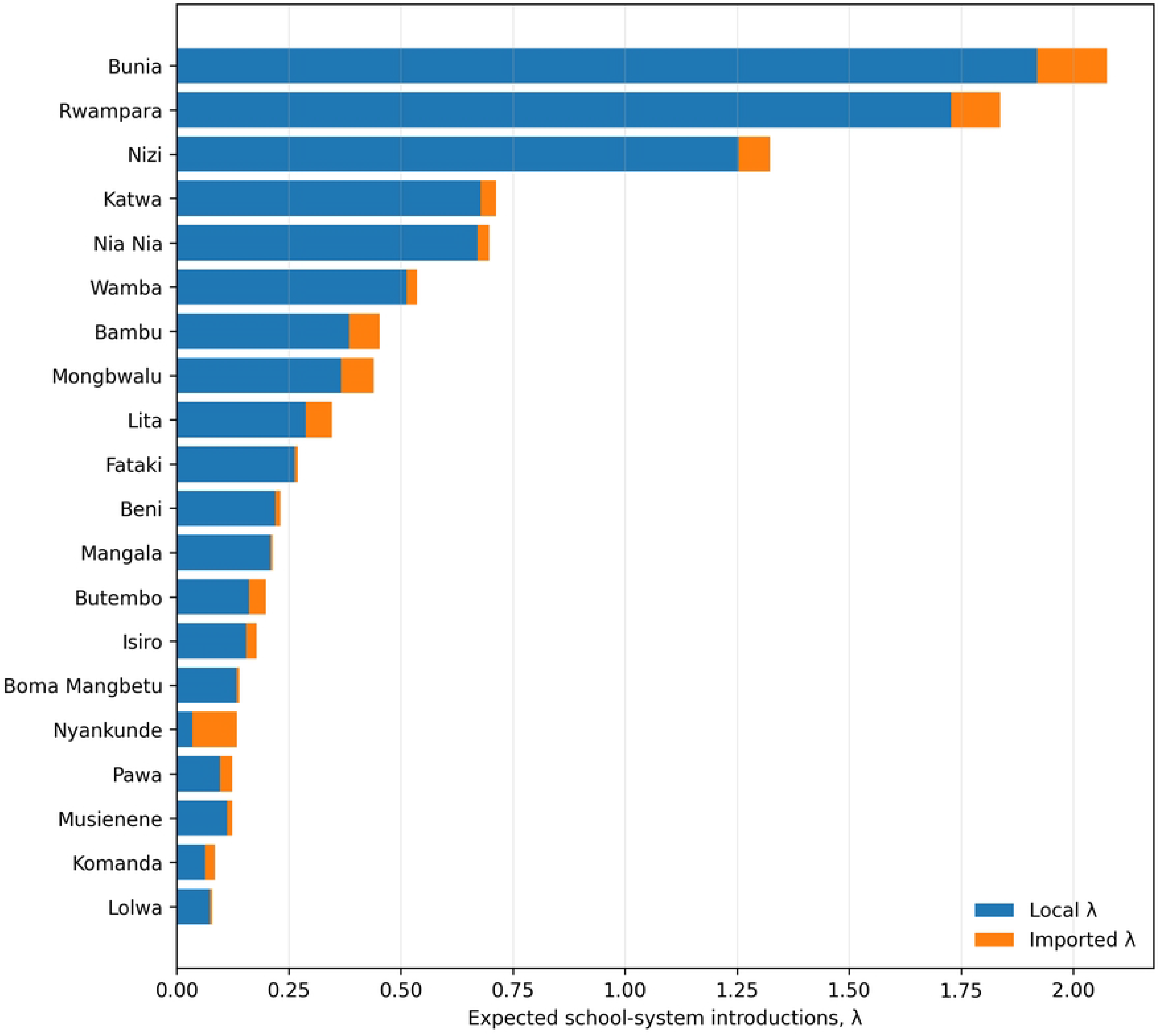
Local and imported components of combined λ among the 20 highest-ranked health zones. Mobility generally broadens the geographic footprint without replacing local epidemic pressure as the dominant component in the highest-risk zones; some health zones, including Nyankunde, have a relatively large imported component.

The effect is particularly clear in places where local incidence alone would be reassuring. In Nyankunde, roughly three quarters of the combined base λ was imported rather than locally generated. At province level, Tshopo had a very small combined absolute expectation, but more than half of that expectation was mobility-derived and 15 health zones had import-only non-zero risk. These examples illustrate why the imported component is best interpreted as a geographic warning signal rather than as an estimate of observed infected travellers.

### Probabilistic sensitivity analysis

Across 10,000 sensitivity draws, the median combined expected introduction count was 11.17, with a 2.5th–97.5th percentile sensitivity range of 5.56–20.97. The width of this interval is important: it shows that the data and assumptions do not support a precise claim such as ‘eleven introductions will occur’. Rather, the analysis supports a broader statement that a non-trivial number of introductions is plausible across the study area under the conditions represented in the model. The median imported component was 1.31, with a corresponding sensitivity range of 0.51–3.28. These are sensitivity intervals, not conventional confidence intervals.

The health-zone ranking was substantially more stable than the absolute count. Bunia, Rwampara, Nizi, Katwa and Nia Nia remained in the top ten in essentially all simulations; Wamba, Bambu and Mongbwalu were also highly stable. Lita remained top-ten in approximately 91% of simulations, while the tenth position was less stable. The main uncertainty was q, the probability that an infected child reaches school before recognition or isolation. Changing the baseline q from 0.10 to 0.50 changed the total expected introduction count from approximately 4.39 to 21.95. By comparison, plausible changes in the mobility scaling moved the total much less.

Put simply, the analysis is more uncertain about whether an infected child will be recognised and intercepted before school attendance than about whether people move between health zones. That is a substantive finding, not merely a statistical one. It directs attention towards contact identification, household reporting, onset-to-alert delay, rapid referral and the practical ability of families and schools to act when symptoms appear.

### Retrospective component validation

Across eight non-overlapping weekly validation origins, the primary analysis included 1,208 health-zone–origin observations, of which 200 showed reported case activity during the following seven days. Recent seven-day local epidemic pressure achieved a pooled ROC-AUC of 0.848. In plain language, when comparing one health zone that subsequently reported activity with one that did not, the local score would rank the subsequently active zone higher roughly 85% of the time. Using a weekly-equivalent 14-day pressure measure increased this discrimination to 0.885, indicating a modest benefit from smoothing recent fluctuations. This interpretation of ROC-AUC describes ranking performance; it is not the probability that any specific health zone will become affected.

Adding mobility produced the largest improvement in geographic discrimination: pooled ROC-AUC rose to 0.952 with seven-day source pressure and 0.953 with fourteen-day pressure. This means that the model became better at placing subsequently active health zones above inactive ones once connectivity was considered. However, the rank correlation with the eventual number of cases was lower after mobility was added. That pattern is coherent: mobility helps identify where some activity may appear, including in connected zones with no recent cases, but it does not tell us how large the subsequent local outbreak will be.

The mobility component was also evaluated directly among health zones that had not yet reported a confirmed case. Across the eight validation origins, 24 such health zones reported a first confirmed case during the subsequent seven-day windows. Using seven-day source pressure, the mobility score achieved a pooled ROC-AUC of 0.876. Thirteen of the 24 newly affected zones (54.2%) were already among the top 10 mobility-ranked unaffected zones and 18 (75.0%) were in the top 20. The median newly affected health zone lay at the 93.5th percentile of the mobility score among zones that were still unaffected at the corresponding cut-off. Operationally, the mobility layer often placed newly affected health zones near the top of the surveillance queue before their first reported case, although it did not identify all of them (Fig 4; Table 3).

**Fig 4.**
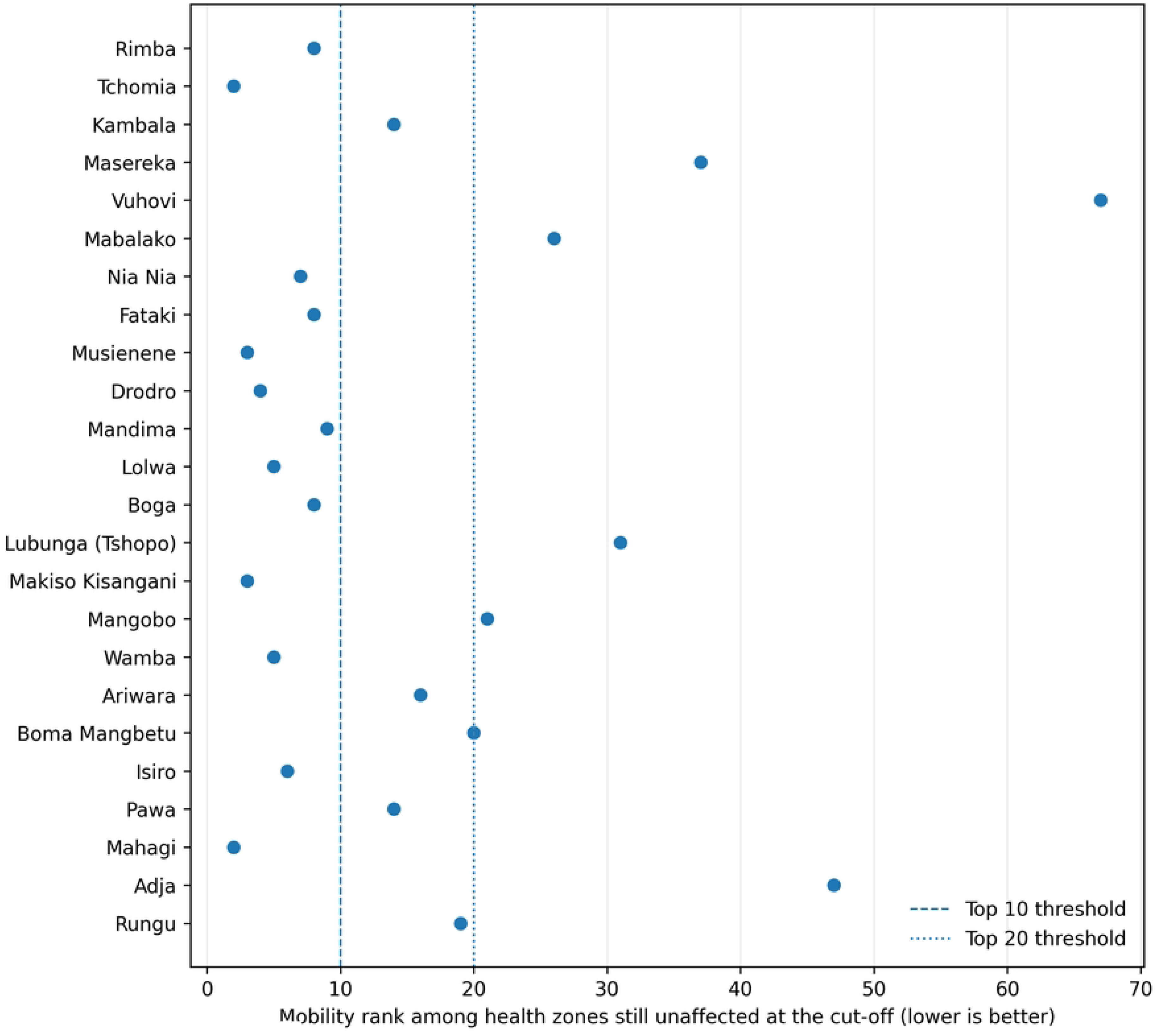
Mobility rank before each first reported health-zone case in the primary seven-day validation windows. Lower ranks indicate higher prior mobility-based concern. Thirteen of 24 first-HZ events were in the top 10 and 18 of 24 in the top 20. Outliers are retained as validation failures; Vuhovi received zero imported risk because no destination flow was represented in the available mobility matrices.

**Table 3.** Retrospective component validation.

note. Primary validation used eight non-overlapping weekly origins: 1, 8, 15, 22 and 29 June and 6, 13 and 20 July 2026.
| Target | Model | ROC-AUC | Average precision | Top-10 capture | Top-20 capture | Interpretation |
| --- | --- | --- | --- | --- | --- | --- |
| Primary: future HZ activity (8 non-overlapping origins) | 7d local | 0.848 | 0.763 | — | 0.669 | Primary short-term geographic ranking. |
| Primary: future HZ activity (8 non-overlapping origins) | 14d local | 0.885 | 0.804 | — | 0.682 | Modest gain from smoothing local pressure. |
| Primary: future HZ activity (8 non-overlapping origins) | 7d mobility | 0.952 | 0.853 | — | 0.661 | Largest gain in geographic ranking. |
| Primary: future HZ activity (8 non-overlapping origins) | 14d mobility | 0.953 | 0.855 | — | 0.677 | Little added gain over 7d mobility. |
| Primary: first case in previously unaffected HZ | 7d mobility | 0.876 | 0.143 | 0.542 | 0.750 | 13/24 first-HZ events in top 10; 18/24 in top 20. |
| Secondary exploratory sensitivity (6 overlapping origins) | Local pressure only | 0.887 | 0.851 | — | 0.546 | Within-sample baseline; not directly comparable with 0.848. |
| Secondary exploratory sensitivity (6 overlapping origins) | Local + surveillance/accesses | 0.886 | 0.849 | — | 0.546 | No measurable incremental gain in the common six-origin sample. |
| Secondary exploratory sensitivity (6 overlapping origins) | Local + surveillance/accesses + mobility | 0.965 | 0.904 | — | 0.540 | Mobility adds discrimination in the common six-origin sample. |

Origin-level sensitivity supported the same qualitative pattern while showing non-trivial between-origin variation. In an origin-block bootstrap over the eight primary origins, mean seven-day local ROC-AUC had a descriptive 2.5–97.5% interval of 0.817–0.885, while mean seven-day mobility ROC-AUC had a corresponding interval of 0.926–0.971. For first-HZ spread, the seven-day mobility ROC-AUC origin-bootstrap interval was 0.834–0.937 and top-20 capture was 0.688–0.969. These intervals reflect variation across a small number of origins and are not an external-validation confidence interval.

Prevalence-aware interpretation strengthens this result. Across the eight seven-day origins there were 930 unaffected candidate health-zone observations and 24 first-HZ events, giving a no-skill AP baseline of 0.0258. The observed AP of 0.143 is therefore about 5.5 times the no-skill prevalence. Random selection would be expected to capture about 2.05 of the 24 events in a top-10 list and 4.10 in a top-20 list, compared with observed captures of 13 and 18, respectively. These are descriptive random-selection baselines, not p-values. Pooled rolling-origin metrics are descriptive because health zones recur across origins; the first-HZ outcome is the direct test of geographic spread.

The misses are informative. Masereka, Vuhovi, Mabalako, Lubunga (Tshopo), Mangobo and Adja fell outside the top 20 before their first reported cases. Vuhovi received a zero mobility score because it was not represented as a destination in the available Flowminder matrix and had no compensating IOM flow. This is a mobility-data coverage failure and illustrates why a low modelled imported risk cannot be treated as proof of no movement or no importation risk.

In the separate six-origin exploratory nested-specification sensitivity, local epidemic pressure alone and local pressure with the available surveillance/access modifiers had almost identical geographic discrimination (pooled ROC-AUC 0.887 and 0.886, respectively), whereas adding mobility increased ROC-AUC to 0.965 and average precision to 0.904. These values should not be compared numerically with the primary eight-origin local-pressure ROC-AUC of 0.848 because the validation samples differ: the primary analysis used eight non-overlapping weekly origins, whereas the nested-specification sensitivity used six overlapping origins concentrated between 13 and 20 July. The comparison therefore supports only an incremental conclusion within the common six-origin sample. It does not mean that surveillance or contact tracing are unimportant. The validation outcome was where cases would subsequently be reported, while q asks a different question: whether an already infected child is intercepted before reaching school. The narrower conclusion is that the available surveillance proxies did not add measurable geographic discrimination in this secondary sensitivity analysis.

Within the same six-origin exploratory sensitivity, the origin-block bootstrap for mean ROC-AUC was 0.866–0.906 for local pressure, 0.864–0.905 for local plus surveillance/access and 0.954–0.974 for the mobility-augmented specification. This reinforces the within-sample ablation result, but it does not create an independent temporal or spatial holdout.

At the frozen 11 August cut-off, the final model ranking was highly robust to the epidemic-pressure window. Replacing seven-day pressure with a weekly-equivalent fourteen-day measure produced a Spearman rank correlation of 0.996; all ten highest-ranked health zones remained in the top ten, and 19 of the top 20 remained in the top 20. Thus, the high-risk cluster is substantially more stable than the absolute introduction counts.

### Recent operational context from SitReps

The recent SitRep synthesis showed that the modelled rentrée risk existed within a response environment that was neither uniformly weak nor uniformly strong. Repeated constraints included incomplete contact follow-up and alert investigation, treatment/referral strain, ambulance and transport limitations, uneven PoE/PoC functionality, insecurity, workforce problems and community resistance. At the same time, the same reports documented expansion of treatment capacity, vehicle and supply deployment, IPC activity and large-scale community engagement [20].

This distinction matters because the numerical introduction estimate and the ability to manage an introduction are not the same quantity. Ituri combined the highest modelled school-introduction pressure with substantial response deployment, yet recent SitReps also documented weak alert investigation on some days, large numbers of unseen contacts in Bunia and Mangala, and heterogeneous treatment and reporting capacity. North Kivu combined lower aggregate modelled pressure with high treatment occupancy, ambulance constraints, insecurity and community incidents. These observations do not validate or invalidate the λ estimates; they describe the environment in which a missed school introduction would have to be detected and managed.

The contextual synthesis also cautions against interpreting low observed incidence as equivalent to strong control. Haut-Uélé had lower case burden than Ituri but weaker contact follow-up and PoE/PoC functionality in some recent reports. Tshopo had low modelled local pressure but also reporting gaps and documented refusal among high-risk contacts. In such settings, ‘low observed pressure’ is a more defensible description than ‘absence of risk’.

No explicit school-specific readiness activity was identified in the verified contextual extracts used for the 31 July–11 August review. This should be interpreted strictly as a reporting observation. It does not demonstrate that school preparation was absent, and generic IPC or community-engagement activity cannot automatically be assumed to have reached schools.

Data coverage was uneven. Of 151 analytical health zones, 96 were flagged as having no health-zone case series and 55 as having an OK/review source status. The Flowminder study matrix had nonzero outgoing rows for 132 health zones and zero outgoing rows for 19; 138 destinations had nonzero incoming Flowminder shares and 13 had none. The IOM matrix was sparser, with nonzero outgoing rows for 43 health zones and nonzero incoming columns for 32. These missingness patterns matter because absent or redacted mobility entries can produce false reassurance, as the Vuhovi validation failure illustrates.

## Discussion

### Principal interpretation

This study addresses a narrow but operationally important question: where is it most plausible that an infected school-age child could reach the school system during the September 2026 rentrée? The answer is geographically concentrated but not confined to health zones with the largest recent case counts. Local epidemic pressure determines most of the highest-risk ordering, while mobility extends non-zero risk into connected zones that may still appear unaffected in routine surveillance. The retrospective analysis supports this geographic use of the framework: recent incidence ranked subsequently active health zones reasonably well, and connectivity substantially improved discrimination.

The framework is therefore stronger as a prioritisation tool than as an exact forecasting tool. The high-risk cluster is robust to alternative recent-incidence windows and to the probabilistic sensitivity analysis, but the total number of introductions is much more uncertain. For non-statistical readers, the distinction is straightforward: the analysis provides a comparatively stable ordering of where attention should be concentrated first, while remaining less certain about the absolute number of infected pupils who may reach school.

### What the validation means—and does not mean

ROC-AUC values above 0.8 are sometimes described simply as good discrimination, but that phrase can obscure what was tested. Here, an AUC of 0.848 for seven-day local pressure means that a subsequently active health zone was generally ranked above an inactive health zone in a retrospective, ascertainment-dependent outcome. When mobility was added, that ordering improved further. The first-case analysis provides an even more tangible result: three quarters of newly affected health zones were already within the top 20 mobility-ranked unaffected zones before their first reported case, substantially above a random-selection benchmark.

This does not validate an exact probability that a child reaches school. The validation outcome is future reported health-zone activity, not unobserved infection, symptom onset, school attendance or within-school exposure. It cannot fully distinguish biological spread from geographic differences in testing access, reporting completeness, insecurity, response attention or data connectivity. The appropriate scientific claim is therefore component-level validation of reported geographic prioritisation: the epidemic-pressure and mobility layers have demonstrated useful discrimination for reported activity, while the school-entry/interception probability remains scenario-based.

### From a health-zone estimate to a real classroom

A health-zone λ only becomes operationally meaningful when translated into the school environment in which an introduction could occur. The literature and contemporary DRC education evidence show substantial variation in classroom conditions. Nationally, rapid enrolment growth following free primary education has intensified pressure on classroom space, while the World Bank continues to identify overcrowding and inadequate water and sanitation as major constraints [8]. In conflict-affected eastern DRC, displacement may add pupils to schools that were already full, convert classrooms or school grounds into shelters, or force education into temporary structures [9,10,12,13].

The urban–rural distinction is useful only if treated carefully. A large urban or peri-urban school may concentrate hundreds of pupils, multiple classes, dense arrival and departure flows and frequent interaction with public transport, markets and surrounding neighbourhoods. A rural school may have fewer pupils but still be operationally vulnerable if water must be fetched from a distance, handwashing stations are absent, classrooms are temporary or damaged, or referral to a health facility requires prolonged travel. Some conflict-affected schools combine the disadvantages of both: displacement creates crowding while insecurity weakens infrastructure and referral. These contrasts are not modelled as a separate urban/rural coefficient because comparable school-level data are not available; they are part of the interpretation of what a health-zone introduction might mean.

This also explains why facilities matter more for containment than for introduction. A handwashing station does not reduce the probability that an infected child exists in the community. It matters once the child reaches school or becomes ill there. Likewise, a temporary isolation space, a trained focal teacher and a functioning referral link do not change community incidence, but they can determine whether a single introduction becomes a controlled referral or a prolonged, confusing event involving classmates, staff and families.

This distinction between health-zone pressure and school-level consequence is central to interpretation. The model assigns an introduction pressure to the school system of a health zone; it does not assume that all schools inside that health zone are equivalent. Two schools facing the same health-zone λ may have very different operational vulnerability because enrolment density, shift systems, water availability, space for temporary separation, staff confidence and referral time differ. The health-zone estimate should therefore be read as the probability of encountering the problem, while school conditions help determine what happens after the encounter. Treating those two levels as separate avoids both false precision and the mistaken conclusion that a health-zone ranking is a ranking of individual schools.

### The central uncertainty is interception before attendance

The largest uncertainty in the model is q, the proportion of infected school-age children who may reach school before they are identified and isolated. This parameter is where community surveillance, contact tracing, household decisions and school behaviour intersect. An exposed child who is already a known and successfully monitored contact may never reach school once symptoms develop. An unrecognised child in a household that does not report early illness may do so. The same recent incidence can therefore generate different school-introduction pressure under different interception conditions.

Ebola contact-tracing literature supports this emphasis: rapid monitoring and isolation of contacts are central to containment [24]. The recent DRC SitReps add operational plausibility by showing that contact follow-up and alert investigation are not uniformly complete. At the same time, our retrospective analysis did not show that the available proxy modifiers improved prediction of where cases would appear next. That is not a contradiction. Geographic spread and pre-school interception are different estimands. The result argues for better direct measurement of q—not for deleting surveillance from the conceptual model or pretending that existing proxies are stronger than they are.

### Mobility and the rentrée as a structured movement event

The rentrée is not only a classroom event. It restarts regular movements between households and schools and changes the timing and density of travel by pupils, teachers and caregivers. Previous work during the 2018–2020 eastern DRC Ebola outbreak showed the value of spatial and mobility models for identifying locations relevant to response placement and importation risk [25]. The 2026 Flowminder data similarly provide a reason not to assign zero risk to a health zone simply because it has not recently reported a case [22,23].

Our validation gives this argument empirical support: mobility improved geographic discrimination and anticipated many first reported health-zone cases. But the failures are equally important. Some newly affected zones ranked poorly because the mobility matrices did not represent their links, and Vuhovi received a zero imported score before its first reported case because no destination flow was available. A modelled zero is therefore a statement about available data, not proof that people do not move.

Mobility should also be interpreted cautiously as a correlated geographic signal. It may partly capture population size, urbanisation, proximity to affected areas, commercial connectivity, reporting intensity or health-system visibility, not only true infectious travel. Because the original rolling-origin score matrices are not fully reconstructed in the submitted package, we did not add a population, distance or urbanisation comparator model. The present result should therefore be read as evidence that the supplied mobility layer improves retrospective reported-activity ranking in this outbreak dataset, not as proof that mobility alone is the causal mechanism.

### Schools can be detection and protection nodes

The Ebola school literature provides an important counterweight to interpretations based only on risk. WHO/UNICEF/CDC guidance and the 2018–2019 eastern DRC experience treated schools as places where illness could be recognised, where referral could be organised, and where teachers and pupils could carry prevention information into households [5–7]. In 2019, thousands of teachers were trained and high-risk schools received handwashing stations, thermometers and hygiene supplies [7]. This history shows that school-health readiness is operationally feasible when it is targeted, simple and linked to the health system.

The lesson is not that thermometers or handwashing stations make schools ‘safe’ in an absolute sense. A school can possess equipment and still fail if teachers fear reporting a child, parents expect punitive exclusion, referral transport does not arrive, or rumours undermine cooperation. Conversely, a modestly equipped school with trusted staff, clear reporting rules and a reliable referral link may function effectively as an early-warning node. Evidence from the 2018–2020 response shows why trust, feedback and adaptation to community perceptions are central rather than peripheral to Ebola control [28–30].

### What minimum school-health readiness means in practice

The literature and the model together support a minimum functional package rather than an unrealistic concept of comprehensive preparedness. The core functions are to recognise an acutely ill pupil or staff member; avoid unnecessary direct contact with body fluids; separate the ill person temporarily without stigmatising them; contact the health-zone response rapidly; clean contaminated areas safely; enable handwashing; and communicate clearly with parents and staff. These functions closely reflect Ebola-specific school guidance, previous DRC practice and current WHO infection-prevention principles [5–7,31].

Feasibility, however, differs by school. The Ministry of Education’s May 2026 prevention instructions—regular handwashing, immediate reporting of suspected cases and reorganisation of space—establish an important national benchmark [15,16]. They should not be interpreted as evidence that implementation is uniform. UNICEF has previously noted that Congolese schools often possess some but not all WASH components needed for effective infection prevention [14], and current education investments continue to target classroom and sanitation deficits [8]. A realistic readiness assessment should therefore ask whether the required function can be performed, not merely whether a national instruction exists.

### Closure, attendance and unintended effects

The analysis should not be read as an argument for blanket closure. The school-closure literature shows that removing children from school can create harms through educational disruption, child labour, early marriage, adolescent pregnancy, exploitation and loss of protective routines [17,18]. In eastern DRC these concerns are not abstract: conflict has already displaced large numbers of children from formal education, damaged schools and increased reliance on temporary learning arrangements [9,10,12,13]. National education indicators also document longstanding urban–rural inequalities in school continuity and basic infrastructure [11].

Closure may be justified temporarily in a specific school or health zone when there is an active case, uncontrolled exposure, inability to refer safely, or another clearly defined trigger. But closure should be treated as an intervention with costs, not as the absence of risk. Children who are not in school remain in households, markets, churches, transport settings, work and informal care networks. The comparative question is therefore which setting offers the better combination of protection, surveillance and continuity under local conditions.

### Why context should qualify rather than re-score risk

The recent SitRep context helps explain why two health zones with similar introduction pressure may face different operational consequences. Treatment-centre saturation, ambulance failure, incomplete alert investigation, non-payment of responders, insecurity or community resistance can slow recognition and referral after an introduction. But these observations were reported unevenly and were not collected as a standardised health-zone survey.

For that reason, converting them into another numerical score would create false precision. A missing SitRep note cannot be interpreted as the absence of a problem, and a repeatedly mentioned problem may partly reflect reporting attention rather than prevalence. Keeping context outside the λ equation preserves an important distinction: the model estimates where introduction is plausible; the contextual synthesis helps interpret what detection and containment may look like if an introduction occurs.

### Strengths and limitations

The main strength of the study is its narrow estimand and transparent separation of processes that are often collapsed into one risk score. Local epidemic pressure, interception before school attendance, imported pressure and post-introduction response are conceptually distinct. The retrospective analysis also tests whether the geographic components add useful information instead of assuming that a more complex model is automatically better.

Several limitations remain. School and pupil denominators are extrapolated rather than drawn from a complete harmonised health-zone school census. The 5–17 case share comes from a national demographic snapshot rather than health-zone-specific age incidence. Mobility data describe population movement rather than observed pupil or teacher travel, and missing or redacted flows can generate false reassurance. Flowminder and IOM channels may partly overlap; treating them as additive may double-count some movement, although a Flowminder-only sensitivity preserved the top-10 ranking and reduced total λ only from 10.974 to 10.736. Contact-follow-up and alert-investigation indicators are affected by reporting completeness. The retrospective validation assesses reported health-zone activity, not the unobserved event of an infected child actually entering a school. The package provides hashes and source lineage for major files but does not constitute an independently double-extracted SitRep audit trail.

The study also does not model within-school secondary transmission. That omission is deliberate. Reliable estimation would require information on the timing and duration of symptomatic attendance, class structure, direct-contact events, body-fluid exposure, cleaning practices, school WASH, isolation and referral. Available evidence is sufficient to identify these mechanisms, but not to calibrate a defensible school-level transmission model across 151 health zones. Non-identifiability is preferable to a false appearance of precision.

### Future empirical refinement

Further empirical refinement should prioritise measurements that replace the largest assumptions rather than additional contextual proxies: age-specific health-zone incidence, known-contact status before confirmation, onset-to-alert and onset-to-isolation intervals, and any documented school attendance or school exposure before detection. Such data would narrow uncertainty around q and the school-age incidence component without changing the conceptual structure.

A later school-level extension would also benefit from administrative data on enrolment, classroom size, shift systems, water availability, latrines, handwashing functionality and referral distance. Such data would be most useful for estimating the consequences of an introduction and the feasibility of minimum readiness, rather than as indiscriminate additions to the primary introduction equation.

## Conclusion

The September 2026 rentrée does not create the underlying BVD epidemic pressure; it reorganises where and when large numbers of children, teachers and caregivers come together. This exploratory framework estimates scenario-based introduction pressure under a frozen 11 August pre-rentrée snapshot. The central epidemiological question is therefore whether an infected school-age child reaches that organised system before community surveillance, household recognition or contact monitoring intervenes.

The model identifies a stable cluster of health zones where that possibility deserves greatest attention. Recent local epidemic pressure explains most of the highest-risk ordering, while mobility materially improves the identification of connected zones that may report their first cases. Retrospective validation supports the geographic prioritisation function of the model, but not a precise forecast of how many infected pupils will attend school.

What happens after an introduction depends on conditions the λ estimate does not measure directly. A crowded urban classroom, a temporary learning space serving displaced families, and a remote rural school without reliable water or rapid referral present different operational problems even when they lie within health zones with similar modelled introduction pressure. This is why minimum school-health readiness—recognition, non-punitive reporting, hand hygiene, safe cleaning, temporary separation and rapid referral—belongs alongside, but not inside, the primary introduction estimate.

The implication is targeted rather than binary. Higher-risk health zones warrant stronger school-linked surveillance and minimum readiness; low observed local incidence should not be treated as zero risk when mobility and reporting gaps remain; and school closure should be considered against realistic educational, protection and community-exposure alternatives. The strongest remaining uncertainty is whether an infected child is identified before school attendance. Improving measurement of that interception process is likely to add more scientific value than further increasing model complexity.

## Data Availability

All data underlying the findings are provided within the manuscript and its Supporting Information. S1 Data FINAL contains the full health-zone model outputs, validation diagnostics, contextual coding, CFR sensitivity, assumptions, exact 10,000-draw PSA national output, deterministic q-only scenarios, validation hardening summaries, missingness/coverage diagnostics and mobility/q details. S2 File FINAL contains compact inputs, exact mobility matrices and study OD shares, exact 10,000-draw PSA output, deterministic q=0.10/0.25/0.50 scenario outputs, source lineage, and Python/R audit and reproduction scripts. The complete historical rolling-origin score-generation pipeline is not fully reconstructed in the compact package.

The underlying public epidemiological and response data are available through the DRC Institut National de Santé Publique SitRep series and the INRB-UMIE BDBV2026-Data repository [19,20]. Derived health-zone outputs, contextual coding, PSA draws, deterministic q scenarios, validation hardening summaries, random-selection baselines, coverage/missingness diagnostics, mobility diagnostics, worked q example and provenance hashes are provided in S1 Data and S2 File. The PSA and q-only calculations are exactly executable from the supplied inputs and code. The full rolling-origin validation score-generation pipeline is not fully reconstructed from dated health-zone histories in the package; instead, the submitted files reproduce validation hardening analyses from supplied origin-level metrics and event ranks. No identifiable patient-level data were used.

## Acknowledgments

Not applicable.

## Supporting information

S1 Data. Supplementary health-zone results, province summaries, validation diagnostics, contextual coding, CFR sensitivity analysis, and model assumptions/provenance.

S2 File. Reproducibility package containing the exact 10,000-draw PSA output, compact health-zone PSA inputs, Flowminder and IOM origin–destination matrices, normalized study OD shares, deterministic q=0.10/0.25/0.50 scenario outputs, origin-level validation summaries, origin-block bootstrap summaries, first-HZ random baselines, missingness/coverage diagnostics, mobility diagnostics, q worked example, source lineage, and Python/R audit/reproduction scripts.

## References

1. World Health Organization. Ebola disease caused by Bundibugyo virus, Democratic Republic of the Congo & Uganda. Disease Outbreak News DON613. 2026 Jul 17. Available from: https://www.who.int/emergencies/disease-outbreak-news/item/2026-DON613

2. Ministère de l’Éducation nationale et Nouvelle Citoyenneté, République démocratique du Congo. Calendrier scolaire 2026–2027 [Internet]. 2026 [cited 2026 Aug 16]. Available from: https://edu-nc.gouv.cd/calendrier-scolaire

3. World Health Organization. What we know about transmission of the Ebola virus among humans. Geneva: WHO; 2014 Oct 6. Available from: https://www.who.int/news/item/06-10-2014-what-we-know-about-transmission-of-the-ebola-virus-among-humans

4. World Health Organization. Ebola disease. Geneva: WHO; 2025 Apr 24. Available from: https://www.who.int/news-room/fact-sheets/detail/ebola-disease

5. World Health Organization, United Nations Children’s Fund, Centers for Disease Control and Prevention. Key messages for safe school operations: in countries with outbreaks of Ebola. Geneva: WHO; 2015 Feb 1. Available from: https://www.who.int/publications/m/item/key-messages-for-safe-school-operations-in-countries-with-outbreaks-of-ebola

6. UNICEF. 80 per cent of school children returned to school in Ebola-affected areas of the Democratic Republic of the Congo. 2018 Oct 12. Available from: https://www.unicef.org/wca/press-releases/80-cent-school-children-returned-school-ebola-affected-areas-democratic-republic

7. UNICEF Democratic Republic of the Congo. Children return to school in Ebola-affected regions of the Democratic Republic of the Congo. 2019 Sep 3. Available from: https://www.unicef.org/drcongo/en/press-releases/children-return-school-ebola-affected-regions

8. World Bank. Democratic Republic of Congo: expanding and equipping primary school classrooms for better learning outcomes. Washington (DC): World Bank; 2024 Feb 26. Available from: https://www.worldbank.org/en/news/press-release/2024/02/28/democratic-republic-of-congo-afe-expanding-and-equipping-primary-school-classrooms-for-better-learning-outcomes

9. UNICEF Democratic Republic of the Congo. The dilemma of schools that shelter DRC’s displaced families. 2023 Sep 2. Available from: https://www.unicef.org/drcongo/en/stories/dilemma-schools-shelter-drcs-displaced-families

10. UNICEF Democratic Republic of the Congo. Temporary learning spaces give displaced children a chance to learn—and make new friends. 2024 Jan 31. Available from: https://www.unicef.org/drcongo/en/stories/temporary-learning-spaces-give-displaced-children-chance-learn

11. UNICEF Democratic Republic of the Congo; Education Cannot Wait. Education Cannot Wait and UNICEF with partners call for substantial increase in funding for crisis-affected children in the Democratic Republic of the Congo. 2022 Oct 26. Available from: https://www.unicef.org/drcongo/en/press-releases/call-substantial-increase-funding-crisis-affected-children

12. UNICEF. Over 130,000 additional children out of school in DR Congo’s Ituri province as violence escalates and aid shrinks. 2025 May 29. Available from: https://www.unicef.org/press-releases/over-130000-additional-children-out-school-dr-congos-ituri-province-violence

13. UNICEF Democratic Republic of the Congo; Education Cannot Wait. Education Cannot Wait launches a new wave of support to strengthen education resilience amid the crisis in Eastern Democratic Republic of the Congo. 2026 Feb 7. Available from: https://www.unicef.org/drcongo/en/press-release/ecw-new-wave-support-strengthen-education-resilience

14. UNICEF Democratic Republic of the Congo. Le Gouvernement du Japon et l’UNICEF appuient la prévention de la COVID-19 dans 2.188 écoles en RDC. 2020 Oct 20. Available from: https://www.unicef.org/drcongo/communiques-presse/gouvernement-japon-prevention-covid-19-ecoles

15. Ministère de l’Éducation nationale et Nouvelle Citoyenneté, République démocratique du Congo. Prévention contre Ebola dans les écoles: le ministère de l’Éducation nationale renforce les mesures sanitaires à l’approche des épreuves certificatives. 2026 May 19. Available from: https://edu-nc.gouv.cd/actualites/prevention-contre-ebola-dans-les-ecoles-le-ministere-de-l-education-nationale-renforce-les-mesures-sanitaires-a-l-approche-des-epreuves-certificatives

16. Ministère de l’Éducation nationale et Nouvelle Citoyenneté, République démocratique du Congo. Ebola: le président de la République ordonne des campagnes de sensibilisation dans les écoles. 2026 May 25. Available from: https://edu-nc.gouv.cd/actualites/ebola-le-president-de-la-republique-ordonne-des-campagnes-de-sensibilisation-dans-les-ecoles

17. Chavez Villegas C, Peirolo S, Rocca M, Ipince A, Bakrania S. Impacts of health-related school closures on child protection outcomes: a review of evidence from past pandemics and epidemics and lessons learned for COVID-19. Int J Educ Dev. 2021;84:102431. doi:10.1016/j.ijedudev.2021.102431.

18. Smith WC. Consequences of school closure on access to education: lessons from the 2013–2016 Ebola pandemic. Int Rev Educ. 2021;67(1-2):53–78. doi:10.1007/s11159-021-09900-2.

19. INRB-UMIE. BDBV2026-Data: data for the 2026 Bundibugyo ebolavirus outbreak [Internet]. GitHub; 2026 [cited 2026 Aug 16]. Available from: https://github.com/INRB-UMIE/BDBV2026-Data

20. Institut National de Santé Publique, Democratic Republic of the Congo. SitRep MVE series, including reports used for the 31 July–11 August 2026 contextual review. Kinshasa: INSP; 2026. Available from: https://insp.cd/

21. UNICEF Data. Democratic Republic of the Congo: demographic and education indicators [Internet]. New York: UNICEF; [cited 2026 Aug 16]. Available from: https://data.unicef.org/country/cod/

22. Flowminder. Population movements to and from affected health zones, based on privacy-secure analysis of mobile operator data from Vodacom Congo, DRC. 2026 Jun 29. Available from: https://www.flowminder.org/resources/publications-reports/drc-reports-publications/population-movements-to-and-from-affected-health-zones-based-on-privacy-secure-analysis-of-mobile-operator-data-from-vodacom-congo-drc-29-june-2026

23. Flowminder. Population movements from the Tshopo health zones, based on privacy-secure analysis of mobile operator data from Vodacom Congo, DRC. 2026 Jul 17. Available from: https://www.flowminder.org/resources/publications-reports/drc-reports-publications/population-movements-from-the-tshopo-health-zones-based-on-privacy-secure-analysis-of-mobile-operator-data-from-vodacom-congo-drc-17-july-2026

24. Webb G, Browne C, Huo X, Seydi O, Seydi M, Magal P. A model of the 2014 Ebola epidemic in West Africa with contact tracing. PLoS Curr. 2015;7:ecurrents.outbreaks.846b2a31ef37018b7d1126a9c8adf22a. doi:10.1371/currents.outbreaks.846b2a31ef37018b7d1126a9c8adf22a.

25. Huber C, Watts A, Thomas-Bachli A, McIntyre E, Tuite A, Khan K, et al. Using spatial and population mobility models to inform outbreak response approaches in the Ebola affected area, Democratic Republic of the Congo, 2018–2020. Spat Spatiotemporal Epidemiol. 2023;44:100558. doi:10.1016/j.sste.2022.100558.

26. Atkins KE, Wenzel NS, Ndeffo-Mbah M, Altice FL, Townsend JP, Galvani AP. Under-reporting and case fatality estimates for emerging epidemics. BMJ. 2015;350:h1115. doi:10.1136/bmj.h1115.

27. Izudi J, Bajunirwe F. Case fatality rate for Ebola disease, 1976–2022: a meta-analysis of global data. J Infect Public Health. 2024;17(1):25–34. doi:10.1016/j.jiph.2023.10.020.

28. Mayhew SH, Kyamusugulwa PM, Kihangi Bindu K, Richards P, Kiyungu C, Balabanova D. Responding to the 2018–2020 Ebola virus outbreak in the Democratic Republic of the Congo: rethinking humanitarian approaches. Risk Manag Healthc Policy. 2021;14:1731–1747. doi:10.2147/RMHP.S219295.

29. McKay G, Baggio O, Camara CA, Erlach E, Robles Dios L, Checchi F, et al. ’The response is like a big ship’: community feedback as a case study of evidence uptake and use in the 2018–2020 Ebola epidemic in the Democratic Republic of the Congo. BMJ Glob Health. 2022;7(2):e005971. doi:10.1136/bmjgh-2021-005971.

30. Earle-Richardson G, Erlach E, Walz V, Baggio O, Kurnit M, Camara CA, et al. New mixed methods approach for monitoring community perceptions of Ebola and response efforts in the Democratic Republic of the Congo. Glob Health Sci Pract. 2021;9(2):332–343. doi:10.9745/GHSP-D-21-00144.

31. World Health Organization. Infection prevention and control guideline for Ebola and Marburg diseases. Geneva: WHO; 2026 May 17. ISBN 978-92-4-011133-2. Available from: https://www.who.int/publications/i/item/9789240111332

